# A pilot clinical study to estimate intracranial pressure utilising cerebral photoplethysmograms in traumatic brain injury patients

**DOI:** 10.1101/2023.05.23.23290325

**Authors:** Maria Roldan, Tomas Ysehak Abay, Christopher Uff, Panayiotis A. Kyriacou

## Abstract

**Objective:** In this research a non-invasive ICP optical sensor has been developed and evaluated in a clinical pilot study. The technology is based on infrared light interrogating brain tissue, including photodetectors used to detect the backscattered light, which is modulated by vascular pulsations in the brain vascular tissue. The hypothesis underpinning this research is based on the expected changes in the extramural arterial pressure affecting the morphology of the recorded optical signals (photoplethysmograms (PPGs)), therefore analysis of the acquired signals using a bespoke algorithm could enable the calculation of the intracranial pressure non-invasively (nICP).

**Methods:** This pilot study is the first evaluation of the nICP probe in patients in whom the gold standard comparator of invasive ICP was available. nICP monitoring was performed for up to 48 hours in each of the 40 patients undergoing invasive ICP monitoring as part of their prescribed medical treatment. The quality of the recorded PPG signals was analysed, and time-based features were extracted off-line. Data from all ICP levels were randomly allocated into two groups to train (80%) and test (20%) a machine learning algorithm. A Bland Altman analysis and ROC curve were calculated to evaluate the accuracy of the estimated nICP value compared to the gold standard of synchronously acquired invasive ICP data.

**Results:** The successful acquisition of cerebral PPG signals from TBI patients for the subsequent extraction of morphological features allowed the generation of a bagging tree model to estimate ICP non-invasively. The non-invasive estimation of ICP was achieved with 95% limits of agreement of 3.8 mmHg and a negligible bias. Furthermore, the model achieved a good correlation coefficient of 82.54% compared to standard clinical invasive ICP monitoring. Finally, the ROC curve analysis showed good diagnostic capability with a sensitivity of 80% and specificity of 89%.

**Conclusion:** Clinical evaluation of this novel optical nICP sensor demonstrated for the first time that it could estimate ICP non-invasively to an acceptable and clinically useful accuracy and paves the way for further technology optimisation and larger clinical studies.

## Introduction

Traumatic Brain Injury (TBI) is a global pandemic affecting 50 million patients globally each year with an annual cost to the global economy of £400 billion^1^. It is the most common cause of death and disability in the under-40 age group, and its incidence is increasing^2^. Secondary brain injury due to raised intracranial pressure (ICP) may result in progressive cerebral ischaemia, herniation syndromes and death at various raised ICP thresholds^3,4^. ICP monitoring is used in unconscious patients to guide treatment^5^, and despite a trial where ICP monitoring was compared to imaging and clinical examination (ICE), which showed no difference in outcomes^6^, ICP-directed therapy remains the international recommended standard of care for severe TBI^7^. A European survey subsequently found that 90% of clinicians would insert an ICP monitor in patients with severe TBI and radiological abnormalities^8^, and a recent meta-analysis suggested that ICP directed therapy was associated with a lower mortality^9^.

All currently available ICP monitoring systems are invasive and require access to the cranial cavity, which have a small but significant risk of complications, including haemorrhage or infection^10^, and for this reason they are generally only inserted by neurosurgeons. The quest for a non-invasive monitor has the immediate advantage that it will eliminate the risk of complications associated with the invasiveness and obviate the requirement for a neurosurgeon to insert it.

Other techniques have been investigated to measure ICP non-invasively^11–15^. These include, Transcranial Doppler (TCD) based techniques^16–21^ or the measurement of the Optic Nerve Sheet Diameter (ONSD)^22,23^ .However, these devices involve cumbersome technology, are operator-dependent, and may not provide direct and continuous measurement of ICP^24^. Also, none of these technologies have been accepted in routine clinical practice for measuring ICP, and hence their clinical efficacy and accuracy has not been extensively demonstrated yet^24^.

In the UK, patients with severe TBI requiring time-critical surgery can take up to 6 hours to reach a neurosurgeon if initially resuscitated in a hospital without a neurosurgery department^25^ and 3.7 hours if taken straight to a neurosurgery hospital^26^. In contrast, a non-invasive ICP monitor used by pre-hospital emergency services would allow ICP-directed therapy (e.g., osmodiuretics, deep sedation and hyperventilation) to be implemented as soon as the patient is reached, certainly within the “golden hour” and at a far earlier stage than what is currently achievable in most situations. Therefore, there is a significant need to develop novel technologies that will allow truly non-invasive and continuous ICP measurements which are neither cumbersome nor operator-dependent.

The technique of near-infrared spectroscopy (NIRS) has been extensively used to assess cerebral oxygenation non-invasively; however, current NIRS devices only provide relative changes of oxy-, deoxy-haemoglobin and tissue oxygenation index (Hb, HbO2, TOI) and only use non-pulsatile brain signals. To-date, there is no evidence of any non-invasive technology that uses the pulsatile component of near-infrared signals to assess ICP quantitatively^14^. Therefore, this research utilised a non-invasive, continuous monitoring system to acquire cerebral pulsatile NIRS signals (Photoplethysmograms (PPGs)), referred as PPG-NIRS, from the forehead of TBI patients. Such technology was developed with the aim of allowing ICP monitoring early and in a variety of settings, thereby decreasing the risks of secondary injury and reducing costs. The developed sensor works by shining infrared light into the brain through the skull, and records PPG signals from the backscattered light detected by two photodetectors.

This pilot study is based on the hypothesis that changes in the extramural arterial pressure (intracranial hypertension) will affect the morphology of the cerebral PPG signals, so advanced algorithms and Machine Learning (ML) models utilising optical signal feature extraction techniques could be implemented in translating the PPG-NIRS signals into absolute measurements of ICP within acceptable accuracy limits. The results of this pilot clinical investigation will guide future device optimisation and the design of subsequent clinical studies.

## Middle section

### Methods

#### Study design

This 78-week, non-randomised pilot study (ClinicalTrials No. NCT05632302) was performed at a single site in the United Kingdom from January 2020 to July 2021 (the study was delayed significantly by the COVID-19 global pandemic). The East of England - Cambridge Central Research Ethics Committee approved the protocol on 14/02/19 (REC reference 18/EE/0276, IRAS ID 219476). The Medicines and Healthcare Products Regulatory Agency (MHRA) had no grounds for objection to making the device available for the purposes of a clinical investigation (CI/2019/0025). This single-arm trial recorded optical signals using the interventional device, while invasive intracranial pressure measurements were recorded simultaneously using a traditional invasive method as part of their normal medical treatment. This study was performed in accordance with the Declaration of Helsinki and in agreement with the International Conference on Harmonisation Guidelines on Good Clinical Practice. This manuscript was written following the preferred reporting items presented on the “*CONSORT 2010 checklist of information to include when reporting a pilot or feasibility trial*” ^27^.

#### Participants

Participants were recruited on the intensive therapy unit (ITU) at the Royal London Hospital, UK on the advice of a personal or professional consultee. Severe TBI diagnoses were made based on guidelines for the Management of Severe Traumatic Brain Injury, Fourth Edition ^7^. With the exemption of one patient, all the subjects were unconscious as a result of head injury, which was the indication of invasive ICP monitoring. One conscious patient undergoing invasive ICP monitoring as an investigation for normal pressure hydrocephalus was also included in the study.

Potential participants were excluded if they were deemed unlikely to survive 48 hours or if a personal consultee advised against participation. Patients who had undergone decompressive craniectomy were excluded due to poor signal quality due to the damping effect of the surgery.

All conscious participants provided written informed consent prior to study participation. In the case patients who could not consent for themselves due to incapacity due to TBI, consent was obtained from a personal consultee or an independent healthcare professional if a personal consultee was unavailable, according to the 2005 UK Mental Capacity Act.

Personal information relating to participants has been kept confidential and managed under the Data Protection Act, NHS Caldecott Principles, The Research Governance Framework for Health and Social Care, and the conditions of Research Ethics Committee Approval.

#### Interventions

The reference invasive ICP monitor used was the Raumedic® Neurovent-P intra-parenchymal pressure probe43 (catalogue reference 092946001) and was used in all patients. This was inserted through a single-lumen 5-French polymer cranial bolt and interfaced with a GE® monitor. The reference invasive ICP was collected via the GE® iCollect software on a laptop computer powered by a medical-grade isolation transformer (SN: 2430).

The developed reflectance non-invasive optical ICP sensor(nICP) was placed on the patient’s forehead, and optical signals were acquired from extracerebral tissues and the brain (Figure 1). The nICP sensor comprised a high-intensity light-emitting diode (OIS-330-810-X, Osaoptolight, Germany) with a peak emission wavelength of 810 nm and two silicone photodiode detectors with a large active area (VBPW34S, Vishay Intertechnology, USA) positioned at 10 mm and 35 mm from the light source^28^. These distances were chosen to guarantee the light penetration necessary to interrogate extracerebral tissues and the cerebral cortex respectively, as is commonly used in cerebral Near Infrared Spectroscopy measurements^29,30^.

**Figure 1:**
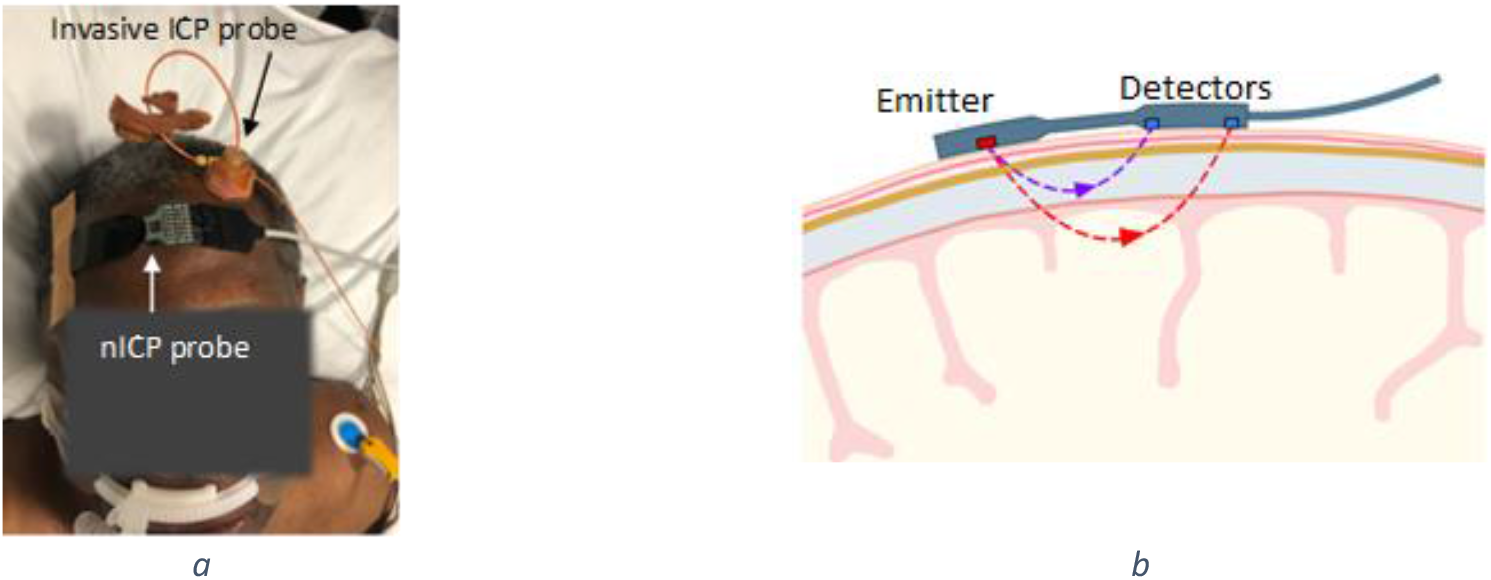
nICP probe: (a) patient with the invasive ICP probe on the left hemisphere and the nICP sensor on the right; (b). The distribution of optical components on the nICP sensor allow the acquisition of deep cerebral PPG-NIRS signals from the distal detector and extracerebral signals from the proximal detector.

The probe was connected to a custom-made instrumentation unit responsible for supplying the driving currents to the LED, transforming and amplifying light intensities into voltages and pre-processing the acquired signals. This system was previously described as the ZenPPG^31^, and it was powered by a battery pack. From the ZenPPG, the signals passed through a National Instruments Data Acquisition Card (DAQ) card to a laptop, where an in-house Labview acquisition software recorded the data sampled at 100Hz.

After the invasive ICP monitor had been inserted, monitoring commenced. The nICP monitor was calibrated in each case, adjusting the LED intensity and amplification gain according to patients’ characteristics and ambient light. Calibration was performed before recording started, then monitoring continued for up to 48 hours, and the investigators monitored the signals during this time. If the patient left the ITU for a scan or surgery, the nICP monitor probe was disconnected but left *in situ*.

### Outcomes

Endpoints, assessment measurements, and the criteria to proceed with a further clinical trial are summarised in Table 1.

**Table 1:** Endpoints, assessment measurements and the criteria to proceed with a further clinical trial

| Endpoint | Assessment measurements | Criteria |
| --- | --- | --- |
| nICP model accuracy | Bland-Altman limits of agreement (LM)<br>Sensitivity (SE) and Specificity (SP) (hypertension threshold = 15 mmHg) | LM ≤ 6 mmHg<br>SE ≥ 80%<br>SP ≥ 90% |
| Quality of the optical signals | Signal-to-noise ratio (SNR) |  |
| nICP probe's safety | Visible skin reaction (VSR)<br>Visible skin burns (VSB) | VSR = mild<br>VSB = none |

The primary endpoint was the accuracy of the nICP model calculated from the acquired optical signals. Moreover, key secondary endpoints were the quality of the optical signals and safety issues related to the probe. Outcomes were evaluated at the end of data collection and complete model analysis for the primary and key secondary endpoints. The nICP model accuracy and the probe’s safety included prespecified criteria used to judge whether or how to proceed with future clinical trials using the nICP monitor.

### Sample size

The GE® monitor uses a sampling frequency of 100 Hz to acquire invasive ICP measurements. Intending to keep the nICP signals consistent with the invasive data, the sampling frequency defined in the LabView interface for the optical signals acquisition was also at 100 Hz. In addition, a monitoring window of up to 48 hours was predefined based on published literature that suggests a range of elective ICP monitoring between 8 hours and 72 hours^32^. Moreover, the selected monitoring window considered that ICP monitoring devices are generally removed once the ICP is normalised with sustained or clinical improvement for at least 48 hours^33^. The features from the optical signals will be explained in the following statistical methods section. Features were extracted from the PPG signals every 60 seconds, leading to a record of up to 2880 observations per patient for each feature.

It remains unclear how to calculate the sample size required for a given machine learning model with the aim of medical application^34^. Therefore, the patients’ sample size was calculated based on a predefined desired nICP model’s diagnostic accuracy. Since the accuracy reported for other non-invasive ICP monitoring techniques^35^ reaches 90% for both sensitivity and specificity, 90% was established as the nICP model’s desired diagnostic accuracy. According to Headway (UK national brain injury charity) statistical reports, the incidence of TBI admission in London hospitals was 216 per 100,000 of the population between 2019 and 2020. Given these assumptions and defining α=0.1 and β=0.2, the sample size was calculated as 37 patients. 10% was added due to potential attrition therefore the final recruitment was 40 patients.

In accordance with the referenced checklist published by ClinicalTrials.gov^36^, this study cannot be considered a small clinical trial despite the intentions of the authors to test a prototype device. Where the primary outcome measure relates to feasibility and not to health outcomes (the latter defined as a trial with at least 10 subjects), the current study would generally not be considered “small” for purposes of the exclusion^36^.

### Randomisation, Implementation and Blinding

Considering that this is a single-arm study, where both nICP and invasive monitoring occurred synchronously in all the patients, there was no intervention randomisation. However, during the off-line analysis of the optical signals, data were randomly assigned to the training or the testing group.

The function *cvpartition* of MATLAB was utilised to randomly allocate the dataset to the training group (80%) and the testing group (20%), following the Pareto law^37^. Because most patients (39/40) were receiving pharmacological sedation with the aim of maintaining an ICP of less than 20mmHg, the distribution of the ICP across the whole dataset is biased toward normal ICP. Therefore, the dataset was divided by ICP levels (5-10mmHg, 10-15 mmHg, …,35-40 mmHg) before applying the partition function to each interval. This ensured that data from all ICP levels were included in both training and testing groups. Due to the nature of the study no blinding was employed. Patients were numbered sequentially by recruitment date using natural numbers starting from number one (1). A key linking these sequential numbers to patient details was stored on a secure clinical computer however other than this, there is no way to track the patient’s personal information.

### Statistical methods

General signal processing was performed using Matlab R2022b (The MathWorks Inc., Natick, MA, USA). All PPG signals are sensitive to different noise sources, such as movement and ambient light. Therefore, movement artefact anomalies and photodetector saturation sections were removed from the cerebral PPG signals, as well the respective synchronous segment was removed from the invasive ICP reference. Then, the cerebral PPG signals were filtered using Butterworth filters to separate the AC PPG component (2nd order bandpass filter with cut-off frequencies of 0.8 and 10 Hz) from the DC PPG component (2nd order lowpass filter with a cut-off frequency of 0.1 Hz). The filtered signals were then normalised by dividing the AC part of the signal by its DC, followed by a 10-factor multiplication.

Several features have been extracted and investigated in the literature to characterise pulsating signals, such as PPG^38^. This study extracted eleven time-domain features from the cerebral PPG signals. These features were:

1. amplitude
2. pulse width
3. rise time
4. decay time
5. upslope
6. downslope
7. area under the curve
8. area of the systolic period
9. area of the diastolic period
10. ratio between both systolic and diastolic areas
11. ratio between the max and min of the second derivative pulse.

The median value of each feature was calculated during a signal window of 60 seconds. Also, the mean value of invasive ICP was calculated for this period. Given the sparsity of recorded pulses above 20 mmHg of ICP, the final dataset only included ICP values between 5 and 40 mmHg. The above procedures resulted in a total of 40,795 observations across all patients.

As mentioned, the dataset of observations (features + mean ICP / window) was randomly portioned in training and testing sets. An additional set (20%) was held out from the training group for cross-validation of the model parameters; this group is called the validation set. Both training and validation sets were used to build a regression bagged tree model using the regression learner app of Matlab R2022b^39^. The functionality and working principle of regression trees and bagging are explained in detail elsewhere ^40^. The regression chosen here was a bagged ensemble of 30 individual decision tree regressors. The maximum depth of each tree was set to 8, considering that an increased tree depth can yield better results but risks overfitting. Also, no restrictions were given on the features available to every node or tree, so all features could be used at any time.

After training the regression forest with the training data set and the corresponding invasively measured ICP as the ground truth, the testing data set was used. Bland-Altman analysis was performed to determine the inter-method agreement. The correlation between methods was assessed by considering the Pearson correlation coefficient. Receiver operating characteristic (ROC) curves were constructed to determine the sensitivity, specificity, and area under the curve (AUC) of the estimated values compared with the measured values for elevated ICP (ICP ≥ 15 mmHg). Statistical significance was defined as p-value < 0.05.

## Results

### Participants’ flow chart and numbers analysed

The participants’ flow diagram shown in Figure 2 includes the number of participants evaluated for potential enrolment into the pilot study and the number excluded at this stage either because they did not meet the inclusion criteria or declined to participate. The diagram also displays information regarding the number of patients included in the main analysis, with numbers and reasons for exclusions. Moreover, the authors included an additional step concerning the random allocation of observations to build the machine-learning model.

**Figure 2:**
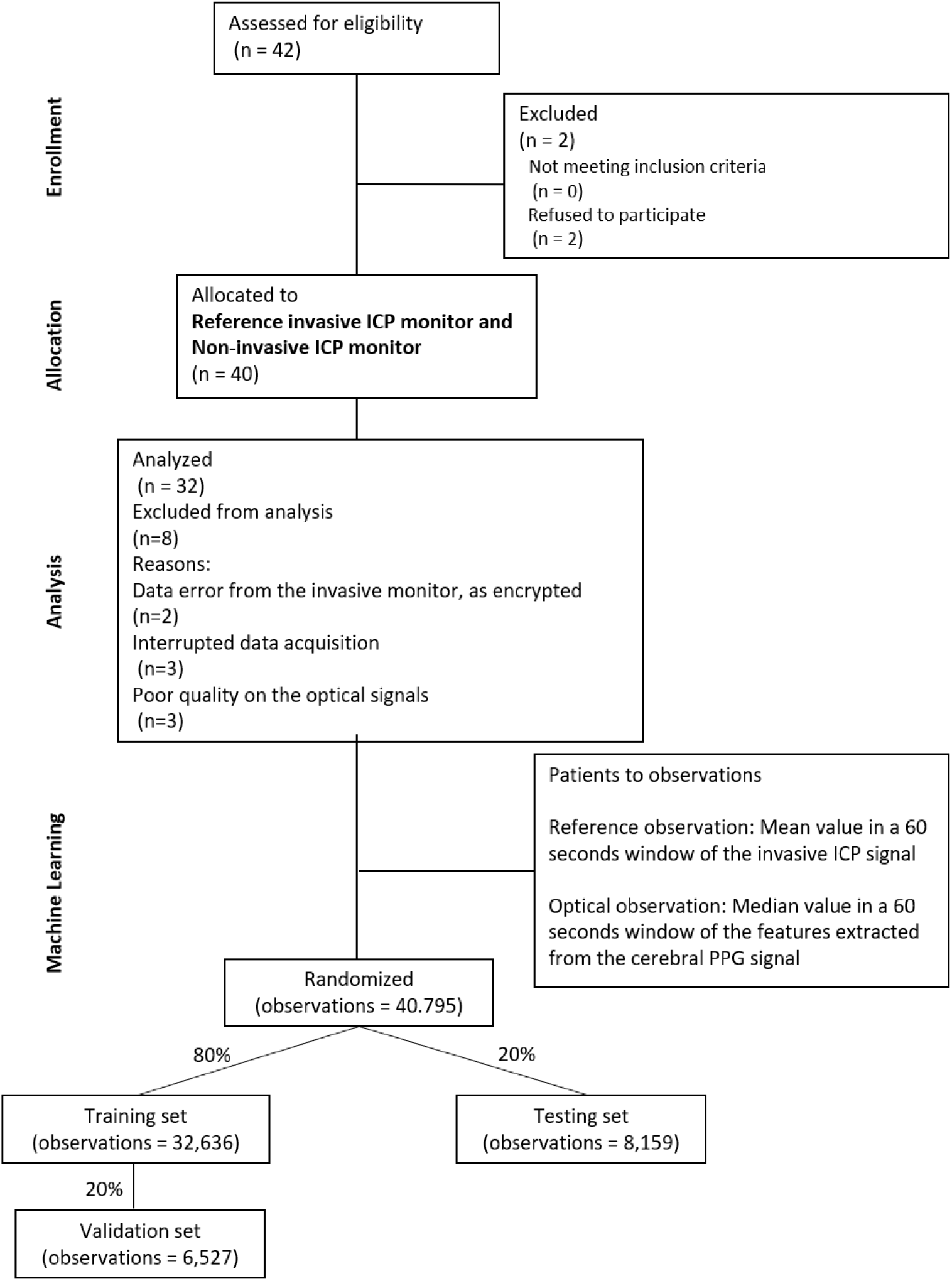
Participants’ flow diagram: the chart shows the enrolment of participants to the study and describe the reasons why some of them were not included in the analysis. In addition to the Consort flow chart, a machine learning data allocation has been included in the figure.

### Recruitment

After ethics approval, 40 patients were enrolled in this single-arm prospective study beginning in January 2020, with completed enrolment in July 2021. All actively participating patients met the inclusion criteria of the study.

### Baseline data

Patient demographics and baseline measures are identified in Table 2. The mean patient age was 42 (SD = 16) years old, with 87% male.

**Table 2:** Baseline characteristics

| Parameter (n=32) | Value |
| --- | --- |
| Age (mean) in years, mean (SD) | 42.16 (16.19) |
| Male, n (%) | 35 (87%) |
| Right probe location, n (%) | 27 (67%) |
| Severe TBI diagnosis*, n (%) | 39 (98%) |

\*One (1) case of possible hydrocephalus
Probe location refers to the hemisphere where the invasive probe was inserted.

### Outcomes and estimation

The distribution of the observations for testing and training was skewed toward lower ICP values (Figure 3a), within the 5 to 40 mmHg range of ICP values. More data were available at lower ICP values, especially between 10 and 15 mmHg, because all patients were undergoing active medical treatment to avoid intracranial hypertension as part of their normal medical treatment. The estimated intracranial pressure (nICP) was compared against the invasive ICP reference on the testing group. Figure 3**Error! Reference source not found**.b shows an uphill pattern from lower to higher ICP values; this indicates a positive relationship between the estimated ICP values and the invasive measurements.

**Figure 3:**
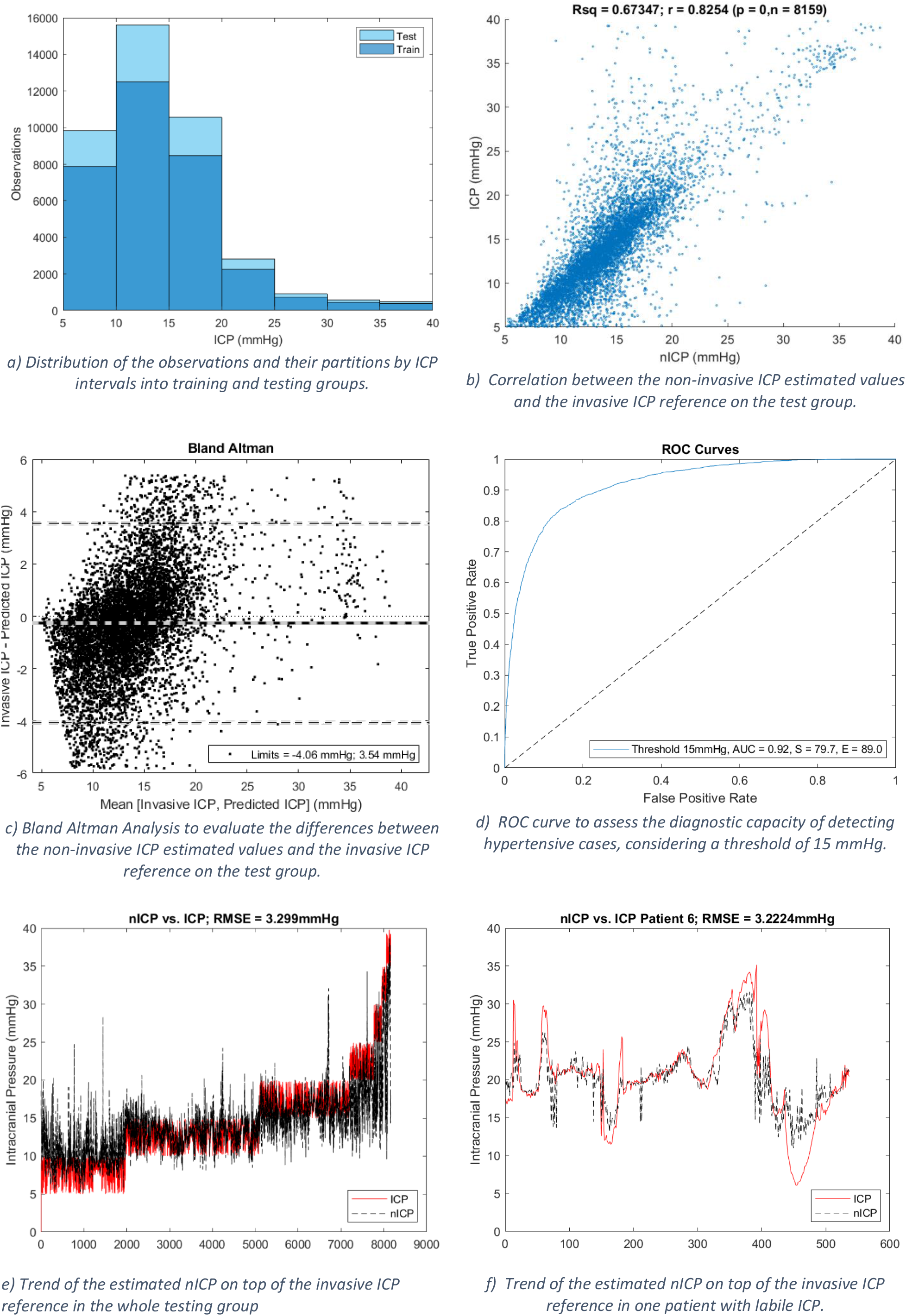
non-invasive ICP model performance

Having a determination coefficient of 0.67 indicates that 67% of the variation in the intracranial pressure is explained by variation in the estimated ICP values. Because the coefficient of determination cannot exceed 100 %, a value of 67 % indicates that the bagging tree matches the actual sample data. Moreover, the estimated and invasive data have a strong correlation of 82.54% (p-value

< 0.05), meaning that an increase in nICP corresponds to an increase in invasive measurement. However, the estimation performance drops for higher ICP values due to lower availability of high ICP data for training the model. Moreover, in order to analyse the agreement between the two methods, a Bland-Altman analysis was performed, comparing the differences in mmHg as shown in Figure 3c. The mean difference, or bias, between nICP and the invasive reference, was of -0.22 mmHg difference. Furthermore, the limits of agreement (Z=1.96) indicate that 95% of the differences between the two monitoring methods are within the range of ±3.8 mmHg. Additionally, the variability of the bias is consistent across the graph, and in accordance with the trend line, the estimated values at low ICP values (<14 mmHg) are higher than the gold standard, while at high ICP (>14 mmHg) are lower. Although an accurate ICP value is useful in the clinical management of TBI patients, the model diagnostic capability to detect hypertensive events could be of interest in neurocritical care. Therefore, Figure 3d presents the ROC curve that evaluated the model performance considering the limitation of an unbalance data distribution within the ICP intervals. Therefore, a lower threshold (15 mmHg) than the one recommended by the clinical guidelines (20 mmHg), was defined. The estimated nICP showed an area under the curve (AUC) of 0.92, a sensitivity of 79.7% and a specificity of 89%.

In addition to the results on the whole dataset, the model was tested individually per patient. Table 3 summarises the mean ± standard deviation and the confidence interval (95%) of the different accuracy parameters evaluated. From this individual analysis, three patients were not included as they did not have more than 100 observations. These results show the variability of the model performance across the patients, presenting reasonable limits of agreement of approximately ± 3.5 mmHg, a mean RMSE of 2.79 ± 0.93 mmHg, and correlation coefficients within 77% and 83% in 95% of the patients.

**Table 3:**
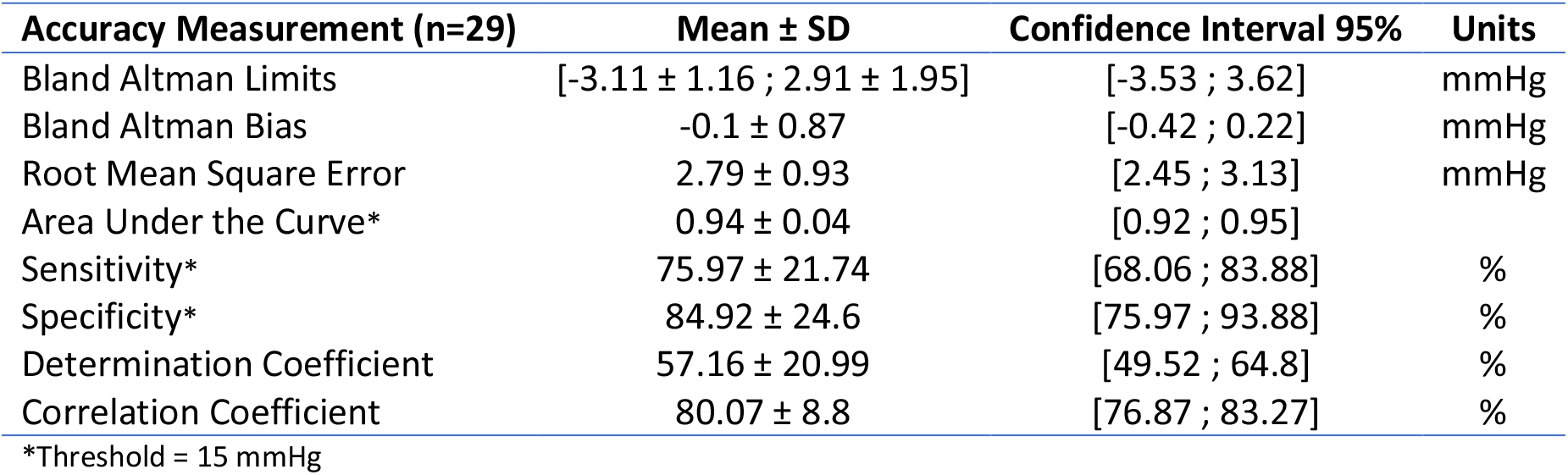
Summary of the model performance at an individual level

Figure 3e shows a continuous estimate of ICP (black line) on top of the testing group’s invasive reference (red). The amount of data per ICP interval is noticed in this figure, as well as the trend of the nICP estimation against the gold standard. Similarly, Figure 3f displays the trending of one random patient who experienced increases in intracranial pressure. Again, the red line matches the trend of the invasive measurements with a root mean square error of 3.22 mmHg.

### Ancillary analyses

The quality of the cerebral PPG signals is key for extracting reliable features from the PPG waveform. Therefore, the signal processing methods included a denoising algorithm with the aim of removing anomalies from the PPG-NIRS signals, as shown in Figure 4. Data length before and after denoising was reduced by 40.18% from 1425.67 hours before denoising to 852.83 hours. However, the Signal Noise Ratio (SNR) increased after the application of the denoising algorithm by 35%.

**Figure 4:**
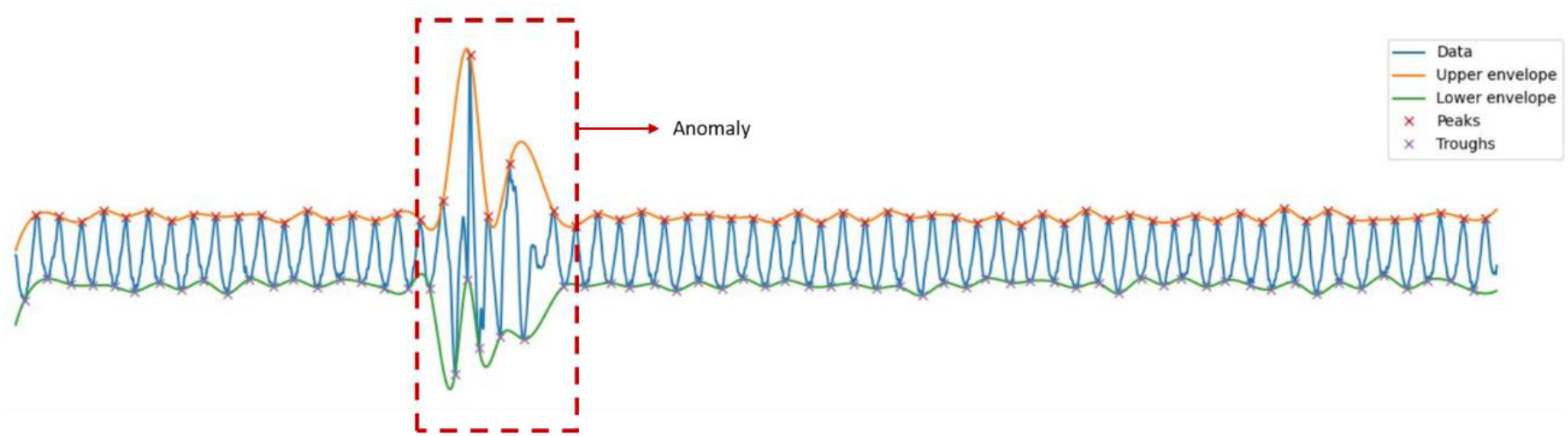
Detection of anomalies by the denoising algorithm: the PPG-NIRS signal in blue colour passes through a denoising algorithm that detect abnormalities as the highlighted by the red square

### Harms

The nICP sensor was attached to the forehead of TBI patients in neurocritical care for up to 48 hours. Near-infrared light was continuously shone into the tissue with a controlled current of up to 30mA. This non-invasive tool did not lead to unintended consequences, harms or effects. Mild marks on the skin were observed after sensor removal, yet no skin lesions or burns were reported. The skin marks were thought to be related to the probe being in place for 48 hours and the marks rapidly disappeared leaving no permanent marks in any participant.

## Discussion

This paper reports a novel optical device’s first clinical pilot study to measure ICP non-invasively using PPG-NIRS waveform analysis. It has several advantages over previously described systems, including TCD and ONSD measurements, in that it is truly non-invasive and is neither operator nor patient dependent.

### Interpretation

The correlation between the invasive and non-invasive instruments observed in this work showed for the first time that the non-invasive ICP monitor effectively followed the trends in ICP changes during the protocol. This provided reassurance that the features extracted from the nICP signals correlate with changes in ICP as initially hypothesised. The estimated nICP also showed good accuracy and good agreement (i.e. RMSE = 3.3 mmHg and limits of agreement of ± 3.8 mmHg) in estimating absolute values of ICP when compared to the reference ICP. Furthermore, the accuracy of different invasive ICP sensors has been reported to be in the range of 0.7-2.3 mmHg^41^, whereas the expected accuracy is ± 2 mmHg for ICP<20 mmHg or ±10% for ICP>20 mmHg^42^. For these reasons, the accuracy between the nICP monitor and the reference invasive ICP sensor observed in this investigation aligned with the expected error for a new ICP measuring technique.

Additionally, this work compares favourably to other non-invasive ICP monitors. For the non-invasive estimation of ICP, a number of TCD-based techniques have been employed. By evaluating a large variety of TCD-based techniques, Cardim and colleagues reported confidence ranges ranging from 4.2 to 59.6 mmHg, with an average confidence interval of about 12 mmHg^20^. Similarly, Bellner et al. looked for the correlation of the TCD pulsatility index with ICP, which led to an accuracy of ± 4 mmHg^43^, yet other studies could not reproduce these results. Kim et al. employed a semi-supervised algorithm to categorise intracranial hypertension by analysing the morphology of the TCD waveform, but this approach could not provide a continuous ICP estimate^44^.

Other recently developed non-invasive techniques either analyse acoustic waves that pass through the skull and provide an ICP estimate accuracy of ±6.8 mmHg ^45^ or measure with ultrasound or other imaging techniques the optic nerve sheath diameter to detect elevated ICP ^46^. Most of the ICP monitoring systems addressed in the review by Rosenberg *et al*. employed CT or ultrasound to assess the diameter of the optical nerve sheath. ^47^. However, they only make a binary decision between elevated and normal ICP, while the nICP sensor reported in this study has the additional advantage of allowing continuous estimations of ICP instead of binary decisions.

Finally, Ruesch *et al*. have been working with NIRS and Diffuse Correlation Spectroscopy (DCS) technologies to estimate ICP non-invasively. The authors present a recompilation of studies where ICP changes were induced incrementally (from about 3-10 mmHg up to 40 mmHg in steps of 10 mmHg) through fluid infusion in non-human primates (n=5 to 8)^48–50^. These studies utilised the cardiac pulse acquired by DCS in a similar way to how this pilot trial used cerebral PPG signals. Interestingly, as explained by Themelis et al., flow and volume are closely correlated^51^. Therefore it is not surprising that morphological features from the flow pulsations change when ICP increases, as the volume signals from NIRS-PPGs do. Nonetheless, Ruesh’s approach relies on incorporating ECG measurements for pulse detection in the DCS signals, which is not needed by the PPG method as these signals are synchronised to the heart cycle. Moreover, Ruesh’s method is not entirely non-invasive, as it utilises MAP measurements from an arteria line to estimate ICP^48–50^.

## Limitations

Assuming that altered cerebral vessels’ geometry brought on by high pressure is the cause of morphological waveform alterations, it is essential to recall that other mechanisms, such as cerebral autoregulation, can also alter the vasomotor tone. The authors accept that cerebral autoregulation is frequently disrupted in patients with intracranial hypertension however this study was not designed to detect this.

An accuracy of ± 3.8 mmHg in the random allocation method may be unrepresentatively good. This is because some data from all subjects are included in the training set. Future work would allow the analysis of additional models and training methods.

In addition, the regression model’s accuracy depends on the training data set distribution. An increasing error with increasing ICP is an effect of fewer training data points at larger ICP values, which is a limitation of the study as most patients are medicated to avoid intracranial hypertension, however this may not be as clinically important as detecting rises in ICP approaching thresholds of 20 or 25 mmHg (the usual cut off for increasing intervention) since anything above this level will be considered for treatment regardless of how far it is above the threshold.

The results presented in this research were analysed off-line however, near real-time waveform analysis would be possible by deploying the algorithms into an embedded system. Future clinical research on an embedded system could provide a performance evaluation of the model in real-time.

## Conclusions

In conclusion, this study has introduced and demonstrated a new and potentially transformative method and an algorithm to estimate ICP in a continuous and truly non-invasive manner which is not co-dependent on any other measurements with cerebral PPG-NIRS. In particular, a bagging tree fed with morphological features from cerebral vessel pulsations that are measured optically by NIRS to estimate ICP values coming from an invasive ICP sensor. The method was demonstrated on adults with TBI.

With further improvements and optimisations of the technology and more validations studies the nICP sensor might be suitable for other conditions including, hydrocephalus, meningitis, and stroke patients, among others. The ability of pre-hospital physicians to initiate ICP-directed neuro-critical care within the “golden hour” (and often several hours before the patient reaches the hospital) may significantly improve outcomes in trauma patients. It may additionally open new scenarios for patients from low- and middle-income countries where the majority of the world’s population has no access to neurosurgery: a low-cost, non-invasive monitor may allow ICP-directed therapy in trauma that could be implemented in any hospital.

## Data Availability

All data produced in the present study are available upon reasonable request to the authors

## Ending section

## Acknowledgements

The authors would like to thank Dr Justin P Phillips for his valuable suggestions on applying cerebral PPG signals for the non-invasive estimation of ICP. Similarly, they want to thank George Bradley for his contributions in signal processing.

## Authorship contribution

M.R: Methodology (supporting); data acquisition (equal); software (equal); formal analysis (lead); investigation (equal); writing – original draft (lead). T.A.Y: Conceptualization (lead); methodology (lead); Resources (lead); software (equal); data acquisition (equal); investigation (equal). C.U: Conceptualization (supporting); Methodology (supporting); project administration (supporting); data acquisition (equal); validation (equal); writing – review and editing (equal). P.K: Conceptualization (supporting); validation (equal); project administration (equal); supervision (lead); writing – review and editing (equal).

## Conflicts of Interest Statement

The authors declare no conflicts of interest.

## Funding Statement

The project is funded by the National Institute for Health Research (NIHR) [Invention for Innovation (i4i) Product Development (Grant Reference Number II-LA-0216-20005)]. The views expressed are those of the authors and not necessarily those of the NIHR or the Department of Health and Social Care.

## Notes

### Competing Interest Statement

The authors have declared no competing interest.

### Clinical Trial

NCT05632302

### Author Declarations

The East of England - Cambridge Central Research Ethics Committee approved the protocol on 14/02/19 (REC reference 18/EE/0276, IRAS ID 219476).

## References

1. Maas AIR, Menon DK, Adelson PD, et al. Traumatic brain injury: integrated approaches to improve prevention, clinical care, and research. Lancet Neurol. 2017;16(12):987–1048. doi:10.1016/S1474-4422(17)30371-X

2. Corrigan JD, Selassie AW, Orman JAL. The epidemiology of traumatic brain injury. J Head Trauma Rehabil. 2010;25(2):72–80. doi:10.1097/HTR.0b013e3181ccc8b4

3. Sorrentino E, Diedler J, Kasprowicz M, et al. Critical thresholds for cerebrovascular reactivity after traumatic brain injury. Neurocrit Care. 2012;16(2):258–266. doi:10.1007/s12028-011-9630-8

4. Åkerlund CA, Donnelly J, Zeiler FA, et al. Impact of duration and magnitude of raised intracranial pressure on outcome after severe traumatic brain injury: A CENTER-TBI high-resolution group study. PLoS One. 2020;15(12):e0243427. doi:10.1371/journal.pone.0243427

5. Smith M. Monitoring intracranial pressure in traumatic brain injury. Anesth Analg. 2008;106(1):240–248. doi:10.1213/01.ane.0000297296.52006.8e

6. Chesnut RM, Temkin N, Carney N, et al. A trial of intracranial-pressure monitoring in traumatic brain injury. N Engl J Med. 2012;367(26):2471–2481. doi:10.1056/NEJMoa1207363

7. Carney N, Totten AM, O’Reilly C, et al. Guidelines for the Management of Severe Traumatic Brain Injury, Fourth Edition. Neurosurgery. 2017;80(1):6–15. doi:10.1227/NEU.0000000000001432

8. Cnossen MC, Huijben JA, van der Jagt M, et al. Variation in monitoring and treatment policies for intracranial hypertension in traumatic brain injury: a survey in 66 neurotrauma centers participating in the CENTER-TBI study. Crit Care. 2017;21(1):233. doi:10.1186/s13054-017-1816-9

9. Shen L, Wang Z, Su Z, et al. Effects of Intracranial Pressure Monitoring on Mortality in Patients with Severe Traumatic Brain Injury: A Meta-Analysis. PLoS One. 2016;11(12):e0168901. doi:10.1371/journal.pone.0168901

10. Tavakoli S, Peitz G, Ares W, Hafeez S, Grandhi R. Complications of invasive intracranial pressure monitoring devices in neurocritical care. Neurosurg Focus. 2017;43(5):E6. doi:10.3171/2017.8.FOCUS17450

11. Khan MN, Shallwani H, Khan MU, Shamim MS. Noninvasive monitoring intracranial pressure - A review of available modalities. Surg Neurol Int. 2017;8:51. doi:10.4103/sni.sni_403_16

12. Zhang X, Medow JE, Iskandar BJ, et al. Invasive and noninvasive means of measuring intracranial pressure: a review. Physiol Meas. 2017;38(8):R143–R182. doi:10.1088/1361-6579/aa7256

13. Roldan M, Abay TY, Kyriacou PA. Non-invasive techniques for multimodal monitoring in Traumatic Brain Injury (TBI): systematic review and meta-analysis. J Neurotrauma. Published online August 21, 2020:neu.2020.7266. doi:10.1089/neu.2020.7266

14. Roldan M, Kyriacou PA. Near-Infrared Spectroscopy (NIRS) in Traumatic Brain Injury (TBI). Sensors. 2021;21(1586):30. doi:10.3390/s21051586

15. Roldan M, Kyriacou PA. Development of a head phantom for the acquisition of pulsatile optical measurements: Mimicking Blood and CSF circulation. [Submitted manuscript]. Published online 2022:15.

16. Brandi G, Béchir M, Sailer S, Haberthür C, Stocker R, Stover JF. Transcranial color-coded duplex sonography allows to assess cerebral perfusion pressure noninvasively following severe traumatic brain injury. Acta Neurochir (Wien). 2010;152(6):965–972. doi:10.1007/s00701-010-0643-4

17. Gura M, Elmaci I, Sari R, Coskun N. Correlation of pulsatility index with intracranial pressure in traumatic brain injury. Turk Neurosurg. 2011;21(2):210–215. doi:10.5137/1019-5149.JTN.3574-10.1

18. Melo JRT, Di Rocco F, Blanot S, et al. Transcranial Doppler can predict intracranial hypertension in children with severe traumatic brain injuries. Child’s Nerv Syst ChNS Off J Int Soc Pediatr Neurosurg. 2011;27(6):979–984. doi:10.1007/s00381-010-1367-8

19. Budohoski KP, Schmidt B, Smielewski P, et al. Non-invasively estimated ICP pulse amplitude strongly correlates with outcome after TBI. Acta Neurochir Suppl. 2012;114:121–125. doi:10.1007/978-3-7091-0956-4_22

20. Cardim D, Robba C, Bohdanowicz M, et al. Non-invasive Monitoring of Intracranial Pressure Using Transcranial Doppler Ultrasonography: Is It Possible? Neurocrit Care. 2016;25(3):473–491. doi:10.1007/s12028-016-0258-6

21. Rasulo FA, Bertuetti R, Robba C, et al. The accuracy of transcranial Doppler in excluding intracranial hypertension following acute brain injury: a multicenter prospective pilot study. Crit Care. 2017;21(1):44. doi:10.1186/s13054-017-1632-2

22. Soliman I, Johnson GGRJ, Gillman LM, et al. New Optic Nerve Sonography Quality Criteria in the Diagnostic Evaluation of Traumatic Brain Injury. Crit Care Res Pract. 2018;2018:3589762. doi:10.1155/2018/3589762

23. Martin M, Lobo D, Bitot V, et al. Prediction of Early Intracranial Hypertension After Severe Traumatic Brain Injury: A Prospective Study. World Neurosurg. 2019;127:e1242–e1248. doi:10.1016/j.wneu.2019.04.121

24. Li Z, Zhang M, Xin Q, et al. Age-related changes in spontaneous oscillations assessed by wavelet transform of cerebral oxygenation and arterial blood pressure signals. J Cereb blood flow Metab Off J Int Soc Cereb Blood Flow Metab. 2013;33(5):692–699. doi:10.1038/jcbfm.2013.4

25. Leach P, Childs C, Evans J, Johnston N, Protheroe R, King A. Transfer times for patients with extradural and subdural haematomas to neurosurgery in Greater Manchester. Br J Neurosurg. 2007;21(1):11–15. doi:10.1080/02688690701210562

26. Bulters D, Belli A. A prospective study of the time to evacuate acute subdural and extradural haematomas. Anaesthesia. 2009;64(3):277–281. doi:10.1111/j.1365-2044.2008.05779.x

27. Eldridge SM, Chan CL, Campbell MJ, et al. CONSORT 2010 statement: Extension to randomised pilot and feasibility trials. BMJ. 2016;355. doi:10.1136/bmj.i5239

28. Roldan M, Kyriacou PA. A non-Invasive Optical Multimodal Photoplethysmography-Near Infrared Spectroscopy Sensor for Measuring Intracranial Pressure and Cerebral Oxygenation in Traumatic Brain Injury. Appl Sci. 2023;13(8). doi:10.3390/app13085211

29. Roldan M., Chatterjee S., Kyriacou P.A. Light-Tissue Interaction Modelling of Human Brain towards the Optical Sensing of Traumatic Brain Injury. In: 43rd Annual International Conference of the IEEE Engineering in Medicine & Biology Society (EMBC). ; 2021:1–4. Accessed January 25, 2022. https://openaccess.city.ac.uk/id/eprint/27326/1/

30. Ferrari M, Quaresima V. Near Infrared Brain and Muscle Oximetry: From the Discovery to Current Applications. J Near Infrared Spectrosc. 2012;20(1):1–14. doi:10.1255/jnirs.973

31. Budidha K, Rybynok V, Kyriacou PA. Design and Development of a Modular, Multichannel Photoplethysmography System. IEEE Trans Instrum Meas. 2018;67(8):1954–1965. Accessed February 7, 2022. https://ieeexplore.ieee.org/stamp/stamp.jsp?tp=&arnumber=8318902

32. Thompson SD, Coutts A, Craven CL, Toma AK, Thorne LW, Watkins LD. Elective ICP monitoring: how long is long enough? Acta Neurochir (Wien). 2017;159(3):485–490. doi:10.1007/s00701-016-3074-z

33. Munakomi S, Das JM. Intracranial Pressure Monitoring. In: StatPearls. StatPearls Publishing; 2022. Accessed November 18, 2022. https://www.ncbi.nlm.nih.gov/books/NBK542298/

34. Balki I, Amirabadi A, Levman J, et al. Sample-Size Determination Methodologies for Machine Learning in Medical Imaging Research: A Systematic Review. Can Assoc Radiol J. 2019;70(4):344–353. doi:10.1016/j.carj.2019.06.002

35. Sallam A, Abdelaal Ahmed Mahmoud M Alkhatip A, Kamel MG, et al. The Diagnostic Accuracy of Noninvasive Methods to Measure the Intracranial Pressure: A Systematic Review and Meta-analysis. Anesth Analg. 2021;132(3):686–695. doi:10.1213/ANE.0000000000005189

36. National Institutes of Health. Checklist for Evaluating Whether a Clinical Trial or Study is an Applicable Clinical Trial (ACT). ClinicalTrials.gov. Published January 18, 2018. Accessed November 18, 2022. https://prsinfo.clinicaltrials.gov/ACT_Checklist.pdf

37. Lipovetsky S. Pareto 80/20 law: derivation via random partitioning. Int J Math Educ Sci Technol. 2009;40(2):271–277. doi:10.1080/00207390802213609

38. Hajj C El. Machine learning techniques for the prediction of systolic and diastolic blood pressure utilising the photoplethysmogram. Published online 2021.

39. The MathWorks Inc. Regression Learner APP. Matlab. https://uk.mathworks.com/help/stats/regression-learner-app.html

40. Sutton CD. Classification and Regression Trees, Bagging, and Boosting. Published online 2005. doi:10.1016/S0169-7161(04)24011-1

41. Zacchetti L, Magnoni S, Di Corte F, Zanier ER, Stocchetti N. Accuracy of intracranial pressure monitoring: systematic review and meta-analysis. Crit Care. 2015;19:420. doi:10.1186/s13054-015-1137-9

42. Kawoos U, McCarron RM, Auker CR, Chavko M. Advances in Intracranial Pressure Monitoring and Its Significance in Managing Traumatic Brain Injury. Int J Mol Sci. 2015;16(12):28979–28997. doi:10.3390/ijms161226146

43. Bellner J, Romner B, Reinstrup P, Kristiansson KA, Ryding E, Brandt L. Transcranial Doppler sonography pulsatility index (PI) reflects intracranial pressure (ICP). Surg Neurol. 2004;62(1):45–51. doi:10.1016/j.surneu.2003.12.007

44. Kim S, Hamilton R, Pineles S, Bergsneider M, Hu X. Noninvasive intracranial hypertension detection utilizing semisupervised learning. IEEE Trans Biomed Eng. 2013;60(4):1126–1133. doi:10.1109/TBME.2012.2227477

45. Ganslandt O, Mourtzoukos S, Stadlbauer A, Sommer B, Rammensee R. Evaluation of a novel noninvasive ICP monitoring device in patients undergoing invasive ICP monitoring: Preliminary results. J Neurosurg. 2018;128(6):1653–1660. doi:10.3171/2016.11.JNS152268

46. Robba C, Santori G, Czosnyka M, et al. Optic nerve sheath diameter measured sonographically as non-invasive estimator of intracranial pressure: a systematic review and meta-analysis. Intensive Care Med. 2018;44(8):1284–1294. doi:10.1007/s00134-018-5305-7

47. Rosenberg JB, Shiloh AL, Savel RH, Eisen LA. Non-invasive methods of estimating intracranial pressure. Neurocrit Care. 2011;15(3):599–608. doi:10.1007/s12028-011-9545-4

48. Ruesch A, Yang J, Schmitt S, Acharya D, Smith MA, Kainerstorfer JM. Estimating intracranial pressure using pulsatile cerebral blood flow measured with diffuse correlation spectroscopy: erratum. Biomed Opt Express. 2020;13(2):710. doi:10.1364/boe.452731

49. Relander FAJ, Ruesch A, Yang J, et al. Using near-infrared spectroscopy and a random forest regressor to estimate intracranial pressure. Neurophotonics. 2022;9(04):1–16. doi:10.1117/1.nph.9.4.045001

50. Tabassum S, Ruesch A, Acharya D, et al. Clinical translation of noninvasive intracranial pressure sensing with diffuse correlation spectroscopy. J Neurosurg. 2022;1(aop):1–10. doi:10.3171/2022.9.JNS221203

51. Themelis G, D’Arceuil H, Diamond SG, et al. Near-infrared spectroscopy measurement of the pulsatile component of cerebral blood flow and volume from arterial oscillations. J Biomed Opt. 2007;12(1):014033. doi:10.1117/1.2710250

